# Evaluation of Rural Public Libraries to Address Telemedicine Inequities

**DOI:** 10.1101/2020.05.27.20113001

**Authors:** Pamela B. DeGuzman, Zack Siegfried, Megan E. Leimkuhler

## Abstract

**Introduction:** In the United States, access to home-based telemedicine is inequitably distributed due to the limited reach of fixed broadband in rural areas. Public libraries typically offer patrons free access to broadband. Libraries, particularly those in rural regions, need to be evaluated as a site for patients to connect to a health care provider over a video visit. The purpose of this research was to evaluate the technological readiness of public libraries to provide telemedicine support and to determine differences in readiness between rural and urban public libraries.

**Methods:** We distributed a survey to members of the Virginia Library Association to evaluate technological readiness of Virginia public libraries to support telemedicine use in their facilities. The survey evaluated each library’s availability and speed of fixed broadband internet access, physical equipment, and private space.

**Results:** Respondents from 39 libraries completed the survey, approximately one-third of which were in rural or small urban areas. All respondents reported fixed broadband, at least four computers, and available staff to assist who technology in their libraries. Eighty-five percent of surveyed libraries reported sufficient broadband speed and a private room available to patrons. There were no significant differences between rural and urban status for any of the library characteristics.

**Discussion:** Public libraries in Virginia are technologically ready to support patrons connecting to health care providers over telemedicine. Systematic guidelines for library-practice collaborations are needed to support implementation across geographic and socioeconomically diverse areas.

## Introduction

Although telemedicine offers tremendous promise to connect rural patients with healthcare providers, access to home-based telemedicine in the United States is inequitably distributed geographically due to the limited reach of fixed broadband (1). The gap between people who have easy access to the internet and those who do not is commonly referred to as the *digital divide* (2,3). The 2019 coronavirus disease (COVID-19) pandemic has increased the urgency behind using technology to deliver health care remotely, particularly for video visits (VVs), a synchronous communication mechanism in which provider assessment is typically conducted using a privacy-enabled videoconferencing platform (4,5). The COVID-19 pandemic has rapidly escalated adoption of these systems (6), but those who live in predominantly rural areas where home-based broadband is either insufficient or unaffordable are likely far less able to connect with providers than their urban counterparts (7).

The ability to deliver VVs to rural populations is stifled by limitations in residents’ accessibility to both the hardware and sufficient broadband speed necessary to stream a videoconferencing call. Several VV platforms use privacy-enabled connections which can be accessed from a cloud-based system or a downloaded application. Smartphones can be used to connect to a VV. While their screen size is far from ideal for connecting to a provider, their use effectively broadens the reach beyond those who own a video-enabled desktop or laptop computer, or tablet (8). At the same time, even smartphones are not ubiquitous. The Pew Research Center estimates that while 95% of U.S. rural residents own a cellphone, only 71% own a smartphone (9). Financial and geographic barriers similarly limit the reach of internet speeds fast enough to support a video call. In the U.S. fixed broadband requires at a minimum 25 megabits per second (Mbps) download and 3 Mbps upload transmission speeds (commonly referred to as “25/3 Mbps”). To successfully conduct to a VV, a fixed terrestrial broadband signal is required; satellite speeds are often too slow, and even in the context of sufficient transmission speed, the several second delay inherent in satellite transmission is highly disruptive throughout a two-way call. Cellular signals often have sufficient video streaming speed, but lengthy video transmission can be highly costly to users (10). Although the availability of fixed broadband signal is increasing, these gains significantly lag in rural areas where perhaps the most generous estimates are that nearly 25% of rural households lack access to fixed broadband (11). In those rural areas where fixed broadband is available, the cost of access is often overly prohibitive to rural residents, who typically have lower incomes (12,13).

### Consideration of Public Libraries as Telemedicine Sites

In light of healthcare organizations transitioning to VV during COVID-19 (6), it seems clear that using telemedicine to provide assessment, education, and treatment recommendations can help maintain care delivery while reducing disease exposure for both patients and providers, regardless of geographic location. However, until fixed broadband access is both geographically and financially accessible, additional solutions are needed to connect residents with health care providers via telemedicine. A systematic evaluation of public libraries as sites from which populations without broadband can connect to VV is needed. Public libraries are a safe space for vulnerable populations (14). They are not only available across the urban-rural spectrum, but also their patrons reflect an aging America. There is an intersection between the age of those with complex health needs and those who visit and utilize internet assets in public libraries (15,16). Libraries are typically visited by older populations seeking health-related information (15). At the same time, older populations are more likely to have multiple chronic conditions (17), requiring care coordination to help manage their complex health needs (16,18). Assisting with access to health information online is already a significant component of libraries’ support of patrons, particularly those post-retirement age (12).

As of the writing of this paper, COVID-19 precautions have closed public libraries across the U.S. (19), likely furthering the digital divide. However, in preparation for communities emerging from social distancing restrictions, public libraries need to be evaluated as places where community members can connect to care providers while minimizing disease exposure. Assessment of public libraries’ *organizational readiness* is an important next step in evaluation of telemedicine implementation (20). Equipment needed for telemedicine VVs includes adequate fixed broadband equipment and speeds; access to video-enabled devices, ideally at least one with a sufficiently large screen for a participant to view a provider (such as a computer or tablet); and a physical space where a participant can speak privately to a provider. The library must also have staff available who are trained in the support of digital technology use. The technological skills needed to assist a patron with a telemedicine connection are typically minimal for a frequent user of internet technology. Similar to connecting to a VV from home, patients are provided with instructions from the provider for connecting to the intervention. Library staff might be called upon to assist to those unfamiliar with how to use a computer devices, or assisting with troubleshooting equipment or connectivity issues.

Once COVID-19 restrictions are eased, healthcare providers need to be ready with strategies to equitably connect populations to telemedicine services, and public libraries should be explored as potential partnering sites. To date, no research has systematically evaluated public libraries’ readiness to provide telemedicine services to rural residents. Thus, the purpose of this research was to evaluate the technological readiness of public libraries to provide telemedicine support. A secondary purpose was to determine if rural public libraries have similar resources to those located in urban settings.

## Methods

We used a quantitative correlational design to address the study aims. We distributed a survey to public librarians and library employees across Virginia to better understand libraries’ readiness to support patients using telemedicine within their facilities, as well as to understand differences between readiness of rural and urban libraries. Similar to many areas of the United States, access to broadband is limited in rural Virginia. Over one-third of rural residents lack homebased access to 25/3 Mbps speed, and 12.9% have no access at all (21). Virginia has at least one public library in every county. **Figure 1** shows the geographic distribution of all public library outlets in Virginia counties against a choropleth map shaded for Census tract-level population of older residents (22).

**Figure 1:**
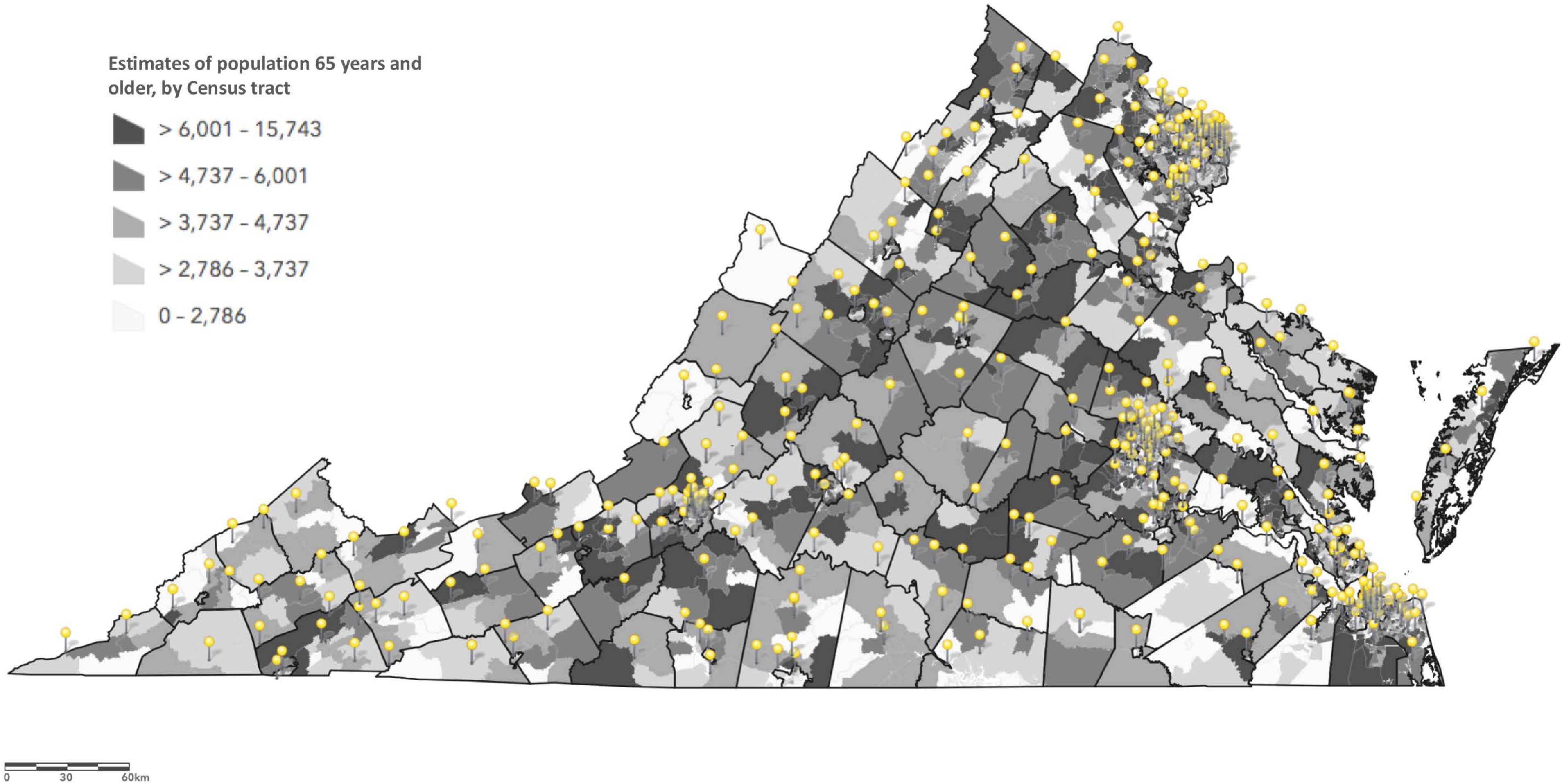
Public libraries in Virginia, indicated by pins. Census tracts are shaded by the percent of the number of people aged 65 or older living in the CT. Map created by PB DeGuzman using ArcGIS Online. Data Sources: *U.S. Public Library Survey 2013 - 2017 - IMSL U.S. Public Library Outlets, Charlie Frye; U.S. Census Bureau's American Community Survey (ACS) 20142018 5-year estimates, Table(s) B01001*

The Institutional Review Board for Social and Behavioral Sciences at the University of Virginia approved the study. The survey was distributed through e-mail to the membership of the Virginia Library Association (VLA). The VLA’s purpose is “to develop, promote, and improve library and information services, library staff, and the profession of librarianship in order to advance literacy and learning and to ensure access to information in the Commonwealth of Virginia,” and its membership includes library professionals (23). The survey was distributed to members using the Qualtrics Survey Tool (Qualtrics, Provo, UT).

We collected information about general characteristics of each library including its physical address, hours of operation and typical number of visits. We additionally incorporated survey questions to evaluate technological readiness including the availability and speed of fixed broadband internet access. Survey questions are listed in **Figure 2**. Of note, we anticipated the possibility that not all respondents would know the precise speed of their library’s broadband, so options were included to indicate the speed if known, and to indicate sufficiency of speed if unknown.

**Figure 2:**
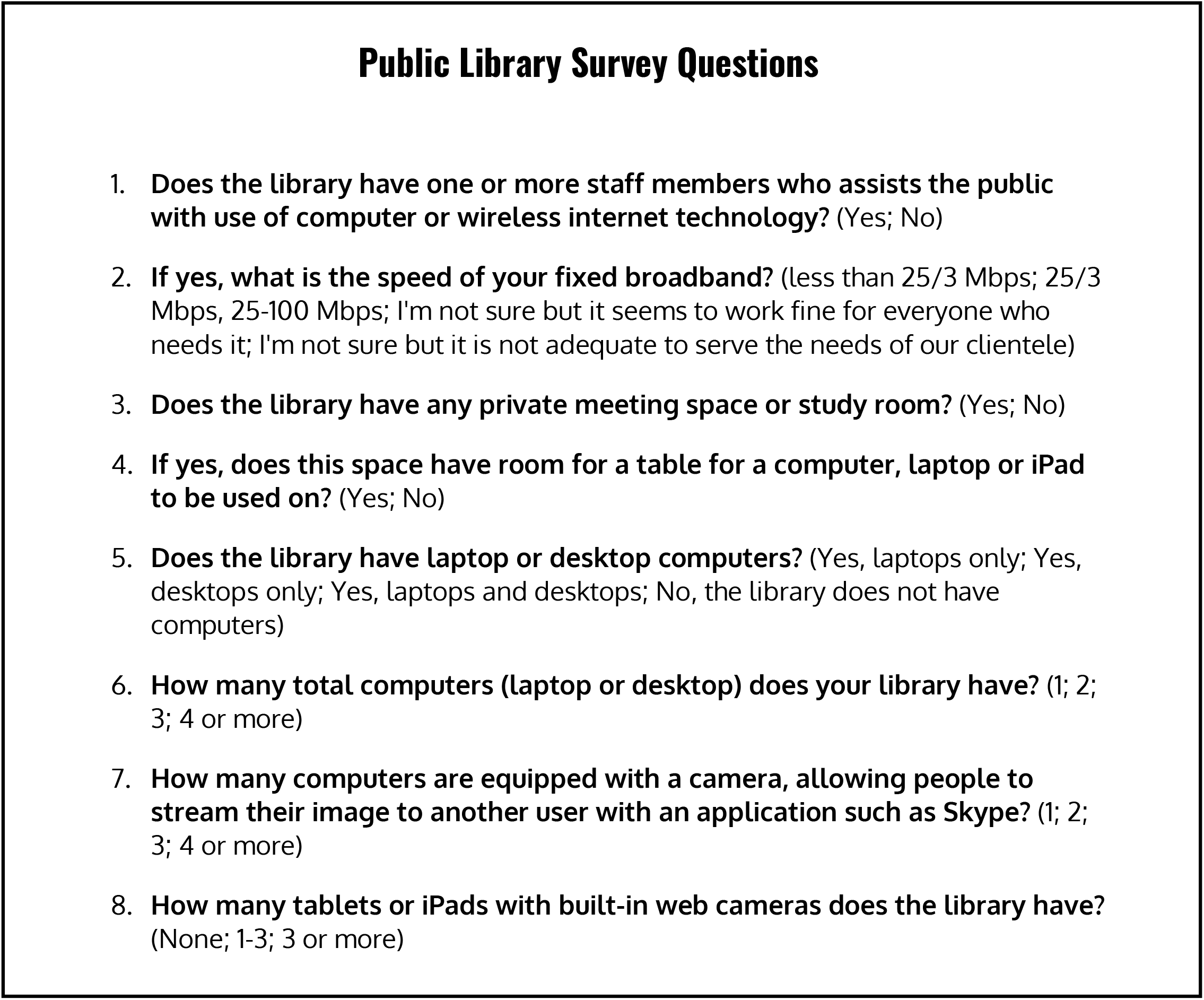
Survey questions distributed to Virginia libraries via the Virginia Library Association membership.

### Data Analysis

Data from public libraries within Virginia were included in the analysis. Location data were used to identify locations on a map and to classify libraries’ rurality using the National Center for Health Statistics (NCHS) Urban- Rural Classification Scheme for Counties (24). Mapping was conducted using ArcGIS Online (Esri; Redlands, CA). Survey data was analyzed descriptively, with some categories collapsed for ease of interpretation. Frequencies were calculated by rurality using NCHS designation status, and chi-square tests for independence were run to determine statistical differences in responses by rurality. Level of significance was set at alpha ≤ 0.05.

## Results

We received surveys from 39 respondents across Virginia. **Figure 3** presents the map of Virginia with the respondent libraries indicated by rurality. Counties are shaded by the NCHS county classification scheme (24). The map shows the broad base of respondents throughout several parts of the state, although there were no respondents from the highly rural southwestern area of the state.

**Figure 3:**
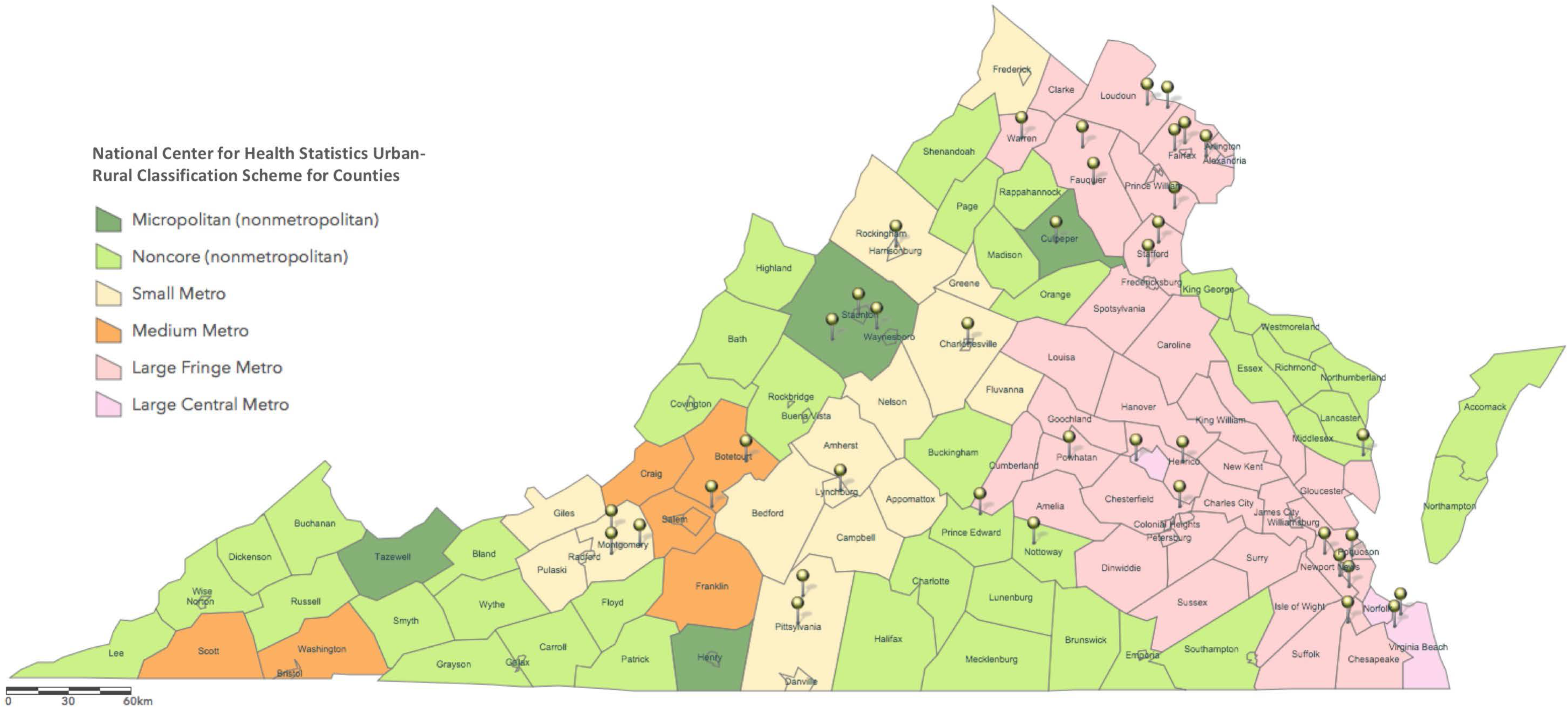
Geographic location of public library survey respondents in Virginia. Respondents indicated by pins. Map is shaded by National Center for Health Statistics (NCHS) Urban-Rural Classification Scheme for Counties. Map created by PB DeGuzman using ArcGIS Online Map created by PB DeGuzman using ArcGIS Online. Data Sources: *Authors’ survey data; NCHS feature layer available via ArcGIS Online*.

For the quantitative analysis, we classified rural as either a 5 (micropolitan) or 6 (noncore) using the NCHS classification; however this yielded only six respondents in the rural category (15%), so we calculated variable frequencies and comparisons with both this traditional classification and a broader definition of rural, additionally classifying category 4 counties (small metropolitan) as rural. Because the small metropolitan counties are adjacent to rural counties, this classification can also reveal useful information. Using the broader classification, 14 libraries (36%) were in or adjacent to rural areas.

The **Table** contains survey responses indicating the technology readiness of our respondents, stratified by rurality status, using both the traditional and broader definitions. All 39 respondent libraries had fixed broadband, at least three computers, and a staff member who could help troubleshoot technology questions. Eighty-five percent of all libraries reported both sufficient broadband speed required to stream a telemedicine videoconferencing application and a private room available to library patrons that would accommodate a computing device. Over half had video-enabled tablets available for patrons. There were no significant differences between rural and urban status for any of the library characteristics, calculated both using the traditional and broad definitions of rural.

## Discussion

Our study suggests that in Virginia, most public libraries have technology necessary to support a VV telemedicine intervention. Rural Virginia libraries do not appear to be different than urban libraries with respect to organizational readiness. However, because individual libraries across Virginia may lack a private space or sufficient broadband speeds to support a VV, individual libraries should be assessed before VV can be pursued. In our sample, 15% of libraries lacked a private space. Lack of space may be due to a small footprint, which is more common for rural libraries; however, the data revealed no differences between rural and urban libraries with respect to private space available. This may be explained by viewing the library through an historic lens. Whereas in the past, public library spaces were designed to support civic, open discourse, the predominant modern function is to be a source of digital information; thus the spatial layout of many of today’s libraries may require adaptation to support current functions (25). Incorporating smaller, private spaces are an important future design consideration; current strategies to overcome this limitation may include patrons connecting from a private vehicle, or temporarily being permitted to use a library office. Most survey respondents reported libraries having either fixed broadband speeds of at least 25/3 Mbps or what they deemed sufficient to support patrons’ use. Still, testing individual libraries broadband to determine its ability to support a VV may be warranted. The U.S. Federal Communication Commission standards suggest that 100 Mbps download speeds are ideal even for smaller libraries (12). Another technological consideration in a readiness analysis is the transportability of computing device assets. Of the libraries in our survey, all owned multiple desktop computers, but far fewer had laptops or video-enabled tablets. In libraries that only own desktops, if these are only available to be used in open spaces, their lack of portability may be problematic for support of private digital conversation. Library acquisition of one or more video-enabled computer tablets should be considered. Tablets have long been ubiquitous in many public school systems due to their low costs (26); thus allocating or raising funds to support additional tablet purchase in a rural county may be a reasonable option.

To our knowledge, this is the first research study to evaluate organizational readiness of public libraries to support VV telemedicine interventions. Important next steps are to evaluate librarians’ perspectives on anticipated barriers and work in collaboratively to develop VV standard protocols that address and overcome these barriers. Librarians already regularly assist patrons with seeking health information online (15), and with training, are open to expanding their roles beyond their foundational training as information specialists (27). Already librarians across the U.S. encounter stark health and social issues including homelessness and opioid overdoses (14,28). While the highly personal nature of these issues may cause librarians to report discomfort with being inadequately trained (29), assisting patrons with connecting to a VV aligns closely with librarians’ traditional roles as information specialists.

### Limitations

The generalizability of our results are limited due to several factors. We used a small, convenience sample of librarians and library staff from Virginia public libraries who self-selected to participate in the survey. The results, including comparison between rural and urban libraries may not be representative of all Virginia libraries or those in other states, and should be interpreted cautiously. We utilized county-level codes to distinguish rural and urban libraries, which may not appropriately reflect differences in library capabilities. The majority of library funding is typically allocated from its local community tax base (12), and county codes do not distinguish between smaller localities that may widely vary by socioeconomic characteristics. Future research evaluating these differences should consider including small area-level socioeconomic status as a factor to identify distinctions in local financial support.

### Conclusion

COVID-19 has brought the social distancing benefit of video visits to the forefront of health care, at the same time as exacerbating the digital divide. As social distancing is eased across the U.S., urgent solutions are needed to ensure those without broadband, most notably rural populations, have equal access to telemedicine. This research study identifies that in many communities, public libraries have the organizational readiness to support telemedicine interventions. Further research to develop systematic guidelines to evaluate and guide library-practice collaborations to implement telemedicine across broad geographic and socioeconomic diverse areas.

## Data Availability

data is available upon request from the corresponding author

## Acknowledgements

*The authors wish to thank Dr. Brian Real at Southern Connecticut State University for his guidance in developing this work*.

---

**Table:**
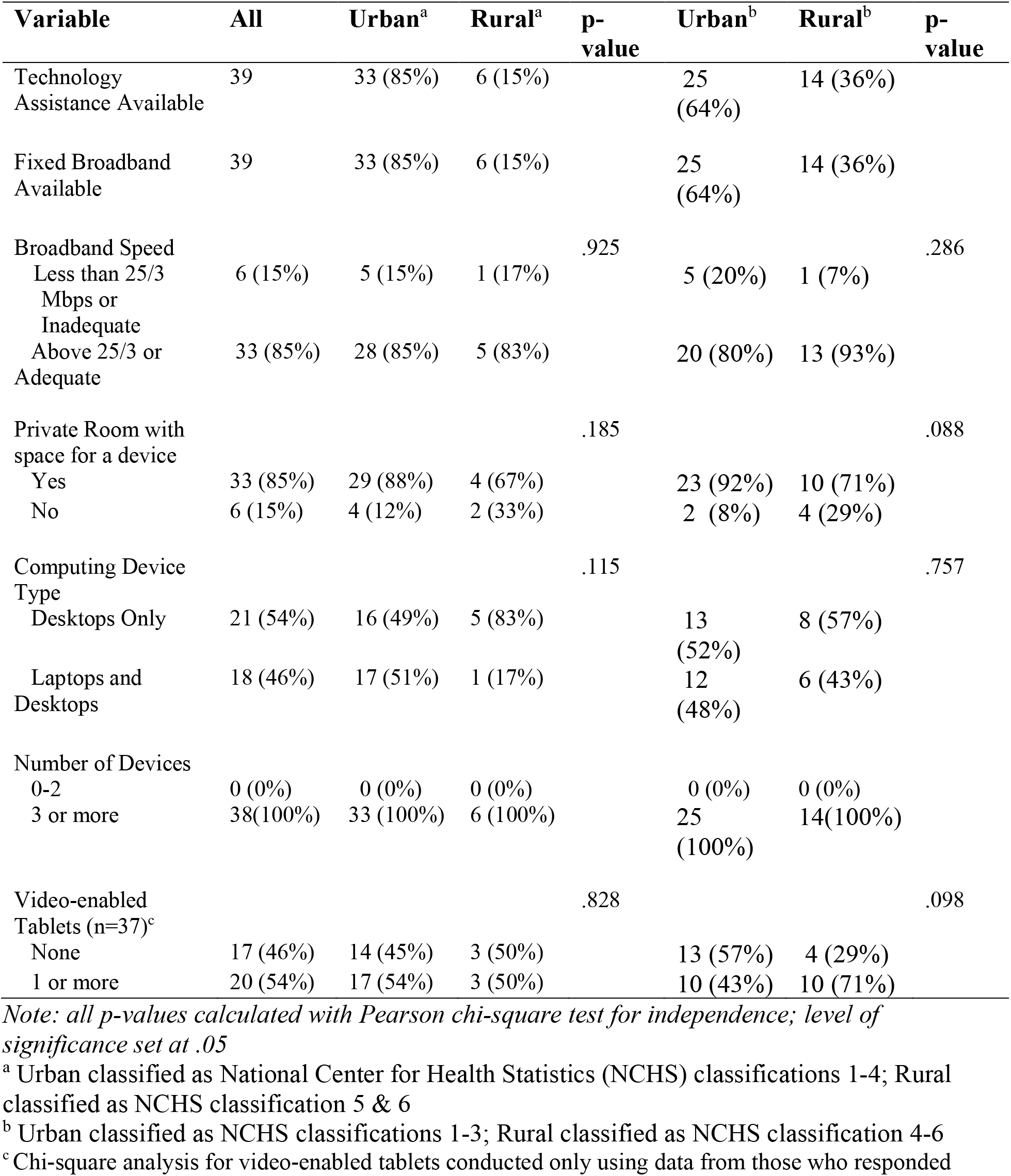
Technology Characteristics of Library Respondents, frequencies (n=39)

| Variable | All | Urban<sup>a</sup> | Rural<sup>a</sup> | p-value | Urban<sup>b</sup> | Rural<sup>b</sup> | p-value |
| --- | --- | --- | --- | --- | --- | --- | --- |
| Technology Assistance Available | 39 | 33 (85%) | 6 (15%) |  | 25 (64%) | 14 (36%) |  |
| Fixed Broadband Available | 39 | 33 (85%) | 6 (15%) |  | 25 (64%) | 14 (36%) |  |
| Broadband Speed |  |  |  | .925 |  |  | .286 |
| Less than 25/3 Mbps or Inadequate | 6 (15%) | 5 (15%) | 1 (17%) |  | 5 (20%) | 1 (7%) |  |
| Above 25/3 or Adequate | 33 (85%) | 28 (85%) | 5 (83%) |  | 20 (80%) | 13 (93%) |  |
| Private Room with space for a device |  |  |  | .185 |  |  | .088 |
| Yes | 33 (85%) | 29 (88%) | 4 (67%) |  | 23 (92%) | 10 (71%) |  |
| No | 6 (15%) | 4 (12%) | 2 (33%) |  | 2 (8%) | 4 (29%) |  |
| Computing Device Type |  |  |  | .115 |  |  | .757 |
| Desktops Only | 21 (54%) | 16 (49%) | 5 (83%) |  | 13 (52%) | 8 (57%) |  |
| Laptops and Desktops | 18 (46%) | 17 (51%) | 1 (17%) |  | 12 (48%) | 6 (43%) |  |
| Number of Devices |  |  |  |  |  |  |  |
| 0-2 | 0 (0%) | 0 (0%) | 0 (0%) |  | 0 (0%) | 0 (0%) |  |
| 3 or more | 38(100%) | 33 (100%) | 6 (100%) |  | 25 (100%) | 14(100%) |  |
| Video-enabled Tablets (n=37)<sup>c</sup> |  |  |  | .828 |  |  | .098 |
| None | 17 (46%) | 14 (45%) | 3 (50%) |  | 13 (57%) | 4 (29%) |  |
| 1 or more | 20 (54%) | 17 (54%) | 3 (50%) |  | 10 (43%) | 10 (71%) |  |

*Note: all p-values calculated with Pearson chi-square test for independence; level of significance set at .05*
<sup>a</sup> Urban classified as National Center for Health Statistics (NCHS) classifications 1-4; Rural classified as NCHS classification 5 & 6
<sup>b</sup> Urban classified as NCHS classifications 1-3; Rural classified as NCHS classification 4-6
<sup>c</sup> Chi-square analysis for video-enabled tablets conducted only using data from those who responded

